# ALPK3 heterozygous truncating variants cause late-onset hypertrophic cardiomyopathy with frequent apical involvement and apical aneurysm

**DOI:** 10.1101/2024.11.14.24317359

**Authors:** Leora Busse, Emily A Huth, Maria Roselle Abraham, Theodore Abraham, Arun Padmanabhan, Julianne Wojciak, Gabrielle Wright, Rajani Aatre, Rachel Campagna, Erika Jackson, Sarah Kreykes, Kimberly Lane, Lindsey Sawyer, Chelsea Stevens, Matthew Thomas, Rebecca VanDyke, Vasanth Vedantham, Emily J Higgs

## Abstract

Hypertrophic cardiomyopathy (HCM) is a genetically heterogeneous disorder with several established genotype-phenotype relationships. While biallelic truncating variants in the *ALPK3* gene cause severe congenital HCM, recent studies have associated heterozygous truncating variants (ALPK3tv) with milder adult-onset HCM. Here we describe a multicenter cohort of 21 individuals with heterozygous ALPK3tv from 10 institutions in the United States, highlighting distinctive clinical characteristics compared to a control group of 132 patients with HCM caused by deleterious variants in sarcomeric genes. As compared to other HCM genotypes, ALPK3tv patients present at an older age (mean 57.25 years) with significantly lower left ventricular wall thickness (14.09 vs 19.78 mm with echocardiogram and 16.13 vs 21.13 mm with cardiac MRI), a lower prevalence of obstructive HCM (15% of ALPK3tv vs 45% of controls), and a strikingly higher incidence of apical aneurysm (22.22% vs. 2.40% in the control group). These results suggest a milder degree of hypertrophy in heterozygous ALPK3-related HCM as compared to other Mendelian causes of HCM, although the increased occurrence of apical aneurysms could have implications for ventricular arrhythmia risk. Our study underscores the importance of recognizing heterozygous ALPK3tv as a cause of adult-onset HCM and provides a comprehensive characterization of its clinical phenotype.

## INTRODUCTION

Hypertrophic cardiomyopathy (HCM), characterized by increased left ventricular wall thickness, affects 1 in 500 to 1 in 200 people across diverse populations ^1^. Individuals with HCM may be asymptomatic with left ventricular hypertrophy (LVH) or present with exercise intolerance, arrhythmia, heart failure, or sudden cardiac death ^2^. A monogenic cause for HCM can be identified in around 40% of HCM patients in well-phenotyped cohorts unselected for family history or age of diagnosis ^3,4^, with the remainder thought to be polygenic or multifactorial ^5,6^. Approximately 30 genes are associated with syndromic and isolated HCM that is inherited in a Mendelian fashion ^7^, most of which are involved in sarcomeric development or function ^8^. *ALPK3* encodes alpha-protein kinase 3, a pseudokinase essential for sarcomere proteostasis. The absence of ALPK3 leads to dysregulation of M-band proteins and impaired sarcomere protein turnover ^9^.

Biallelic truncating variants in *ALPK3* were established in 2016 to cause severe congenital or pediatric-onset hypertrophic or dilated cardiomyopathy ^10^. Several reports since then have expanded the phenotype of this recessive condition to include variable expression of facial dysmorphism and skeletal abnormalities ^11–13^. Some of the pedigrees ascertained via the bi- allelic pediatric probands revealed heterozygous relatives with adult-onset cardiomyopathy ^10,12,13^ leading to the hypothesis that monoallelic loss of function of *ALPK3* cause an adult-onset phenotype while bi-allelic loss of function of *ALPK3* cause severe pediatric-onset phenotype.

More recently, several cohort studies have added to the evidence that individuals harboring a deleterious *ALPK3* variant in heterozygosity may develop HCM ^13–17^.

While several publications have described aspects of the phenotype of *ALPK3* truncating variant (ALPK3tv) heterozygotes, there have been conflicting data with respect to age of onset and severity of phenotype in comparison to other monogenic causes of HCM. In addition, a higher frequency of apical involvement has been observed in some other cohorts but whether this is associated with other phenotypes including apical aneurysm has not been defined, nor have MRI findings been characterized in these patients. Here we report the first multicenter cohort of ALPK3tv heterozygotes from 10 centers in the United States. We provide a comprehensive description of the clinical characteristics of ALPK3tv heterozygotes with HCM compared to patients with HCM caused by sarcomeric genes, including 3 generation pedigrees, MRI findings, frequency of arrhythmias, and frequency of left atrial abnormality. Taken together with earlier studies, these data provide a fuller picture of the clinical spectrum of ALPK3tv heterozygous disease with implications for management of affected individuals and for asymptomatic carriers.

## METHODS

### Identification of ALPK3 Heterozygotes

We invited members of the National Society of Genetic Counselors’ Cardiovascular Special Interest Group (SIG) via the SIG listserv to collaborate. The invitation asked members of the SIG to identify affected individuals with a heterozygous *ALPK3* variant on clinical genetic testing. De-identified case data was gathered by collaborators via chart review and submitted to the research team at the University of California San Francisco (UCSF) via a secure REDCap form. The data fields gathered for each case were pre-defined by a group of cardiovascular genetic counselors, cardiologists and researchers at UCSF and included history of hypertension, presence of comorbidities, symptoms at diagnosis, phenotypic features, treatment, most recent cardiac imaging results (or most recent pre-treatment results for individuals who underwent myectomy or alcohol septal ablation), presence of arrhythmias, electrocardiogram (ECG) data, genetic test results and a 3 generation pedigree. Self-reported ancestry was not collected as it was deemed to have limited utility in this context; although shared genetic or environmental modifiers might influence HCM presentation, self-reported ancestry is an unreliable proxy for these factors ^18^.

### Inclusion and Exclusion Criteria

Patients were included if they were heterozygous for an *ALPK3* variant reported by the clinical genetic testing laboratory as pathogenic or likely pathogenic and had a clinical diagnosis of cardiomyopathy as documented by the genetics clinic involved in their care. Patients were excluded if genetic testing revealed another potential genetic cause, if they had ALPK3tv but the clinical genetic testing lab did not classify it as pathogenic or likely pathogenic, or if they did not have cardiomyopathy (familial variant testing or genetic testing for other reason).

Statistical analyses were performed using R Studio. Continuous variables were compared using two-way t-tests or Wilcox tests as appropriate, and categorical variables were compared using two-tailed Fisher’s exact tests. This study was approved by the UCSF Institutional Review Board (IRB # 23-40474); no informed consent from participants was required.

## Results

### Characterization of ALPK3tv heterozygote cohort

Nine submitted cases were excluded from analysis as they did not meet the inclusion criteria. One case was excluded because the individual carried a likely pathogenic *MYBPC3* variant in addition to a pathogenic *ALPK3* variant. Six cases were excluded because although they carried pathogenic ALPK3tv they did not have cardiomyopathy; 5 of these are genotype-positive phenotype-negative relatives of ALPK3tv probands where we did not have access to the proband’s clinical information, and 1 had genetic testing in the setting of cardiac arrest but had no left ventricular hypertrophy. One case was excluded because although they had an ALPK3tv, the clinical genetic testing laboratory classified the variant as uncertain significance. These cases, while excluded from statistical analysis, are included in the Supplemental table 1.

Our final *ALPK3* cohort was composed of 21 individuals with a heterozygous pathogenic or likely pathogenic *ALPK3tv* from 20 families. The majority of cases (76%) were contributed from multidisciplinary cardiovascular genetics clinics at hospitals affiliated with HCM Centers of Excellence, and the remainder were contributed from multidisciplinary cardiovascular genetics clinics at other hospitals (14%), and medical genetics clinics (10%). A clinical diagnosis of HCM was present in 20 cases (95%), and 1 had a diagnosis of left ventricular non-compaction with LVH. The genotypic and phenotypic characteristics of these 21 individuals are summarized in Tables 1 and 2, respectively. The individuals in our cohort underwent genetic testing between 2016 and 2024. Almost all received genetic testing from Invitae (18/21, 85.7%), with others receiving testing from GeneDx (2/21), and Ambry Genetics (1/21). All individuals had comprehensive clinical diagnostic genetic testing, other than one affected individual who had familial variant testing as there was already a known ALPK3tv in their family. Approximately half of individuals (11/21, 52.4%) received a combined cardiomyopathy and arrhythmia panel, a third (7/21) received a HCM panel, and 2 (9.5%) received a cardiomyopathy panel.

**Table 1.**
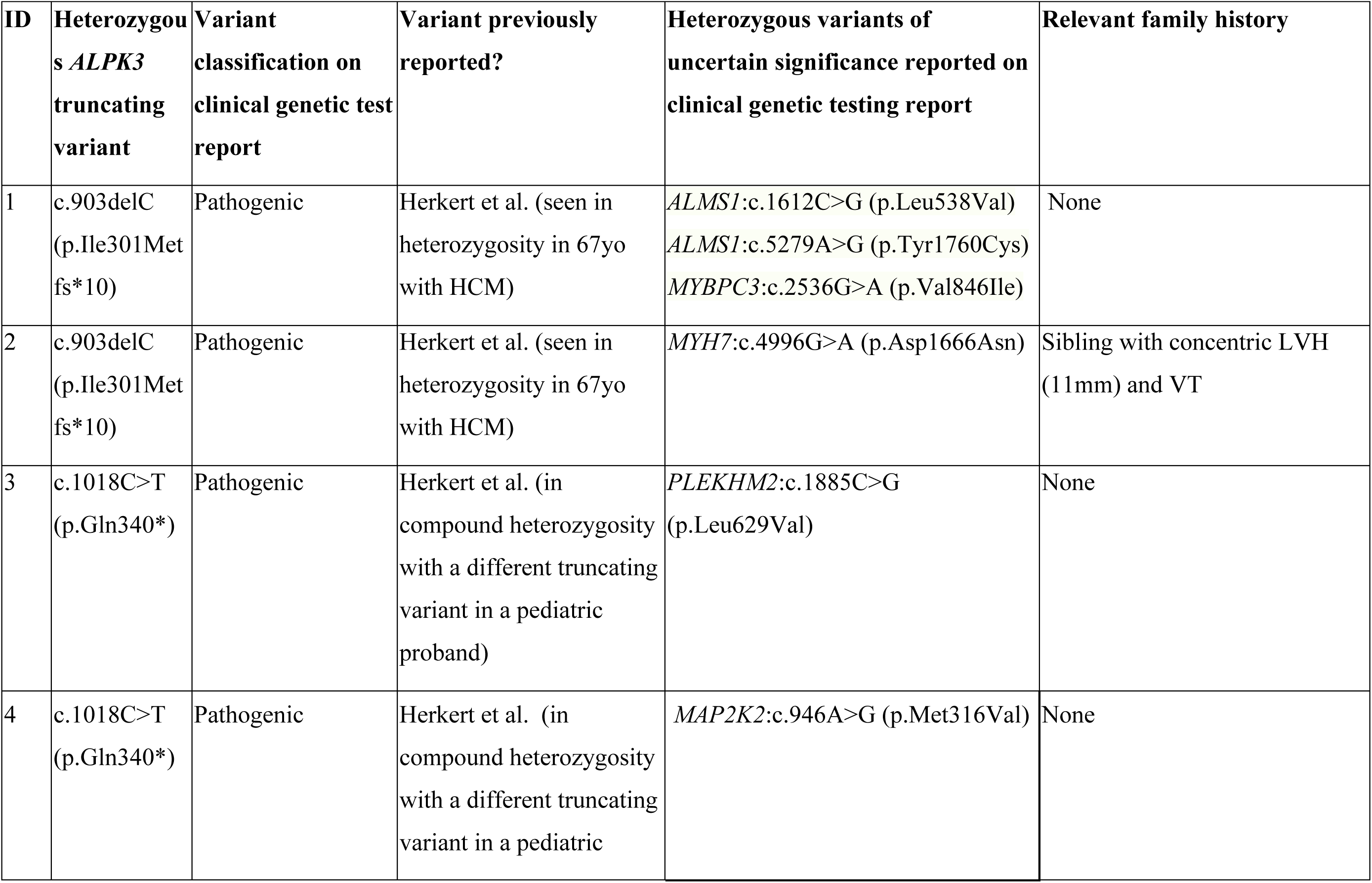
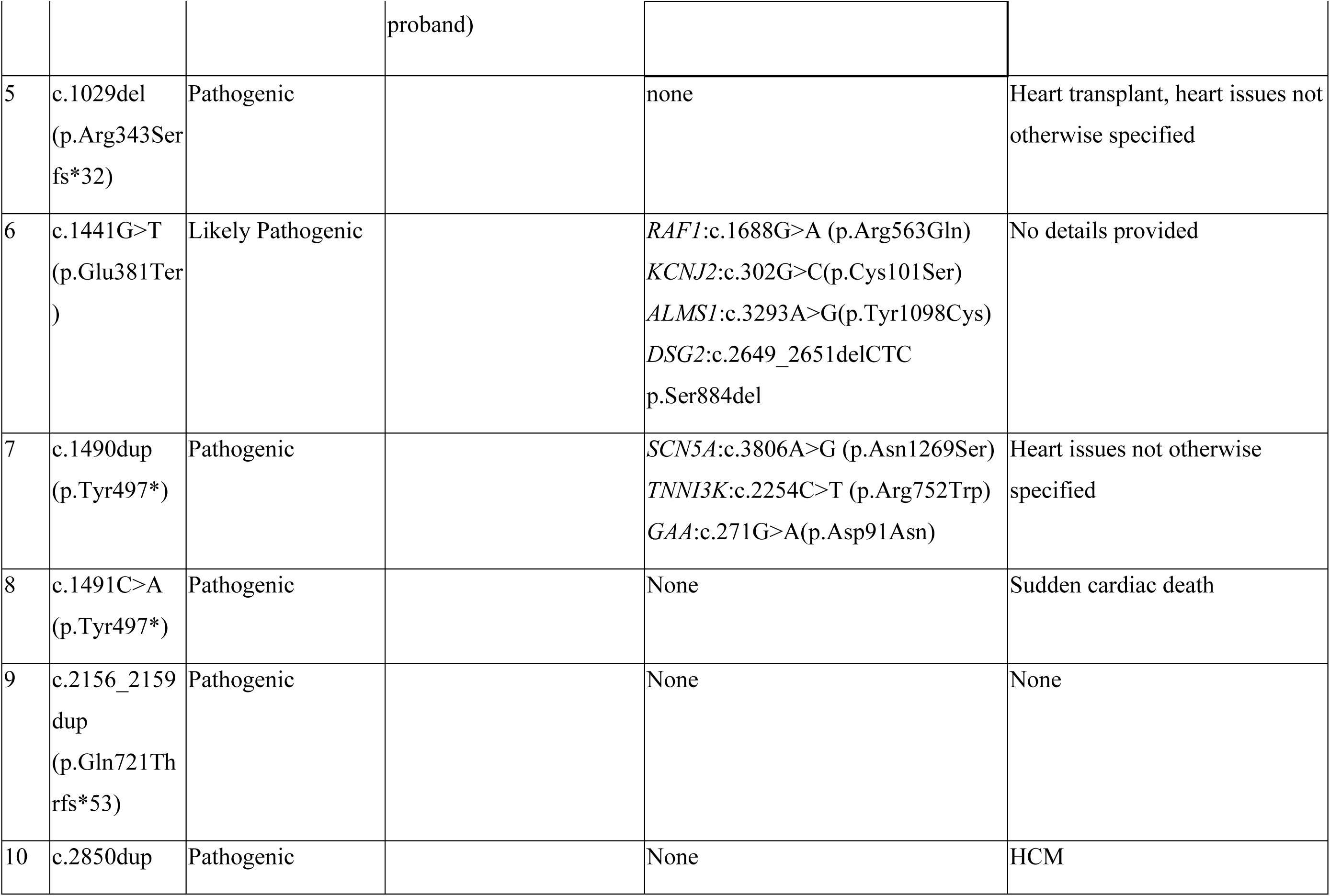
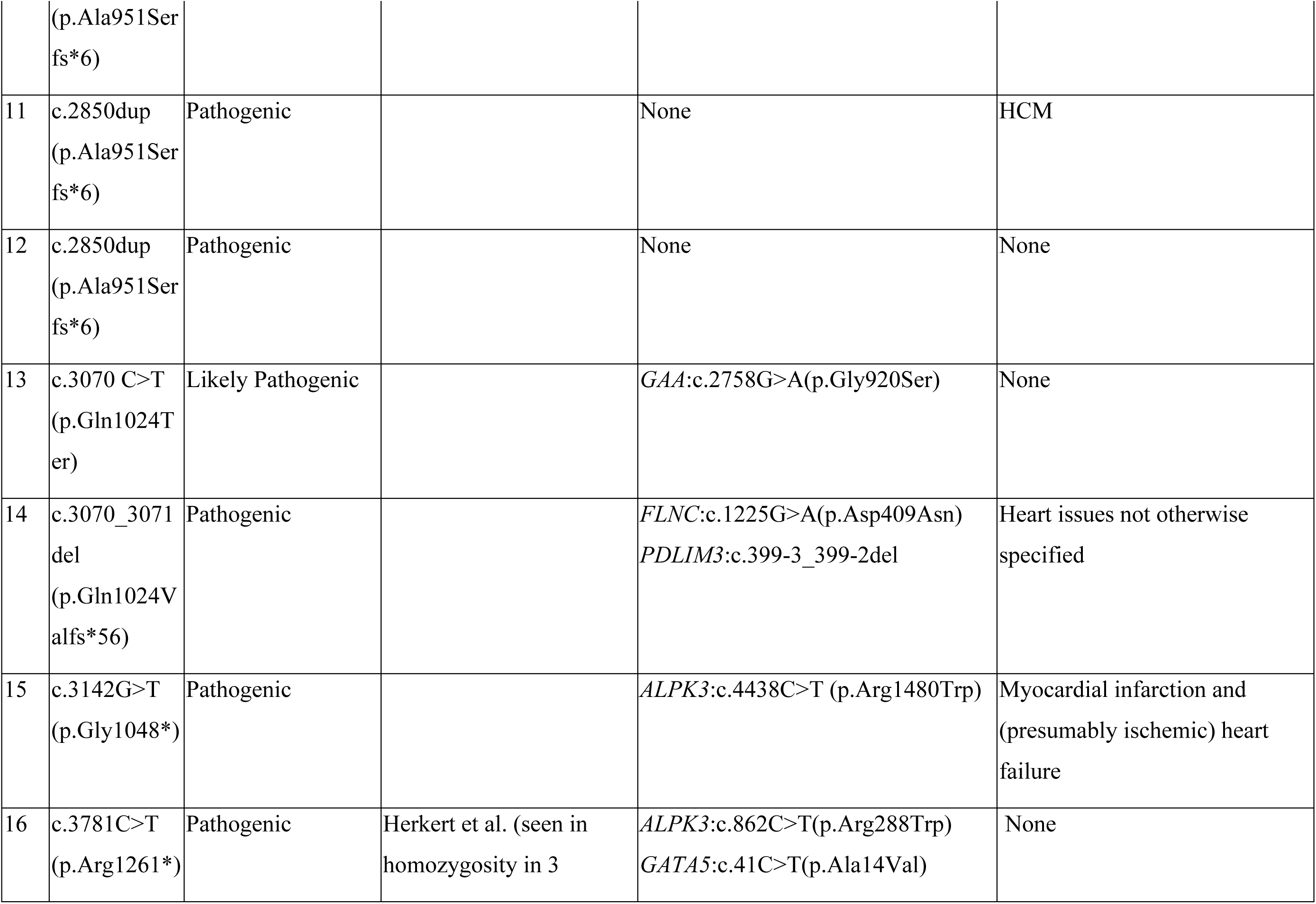
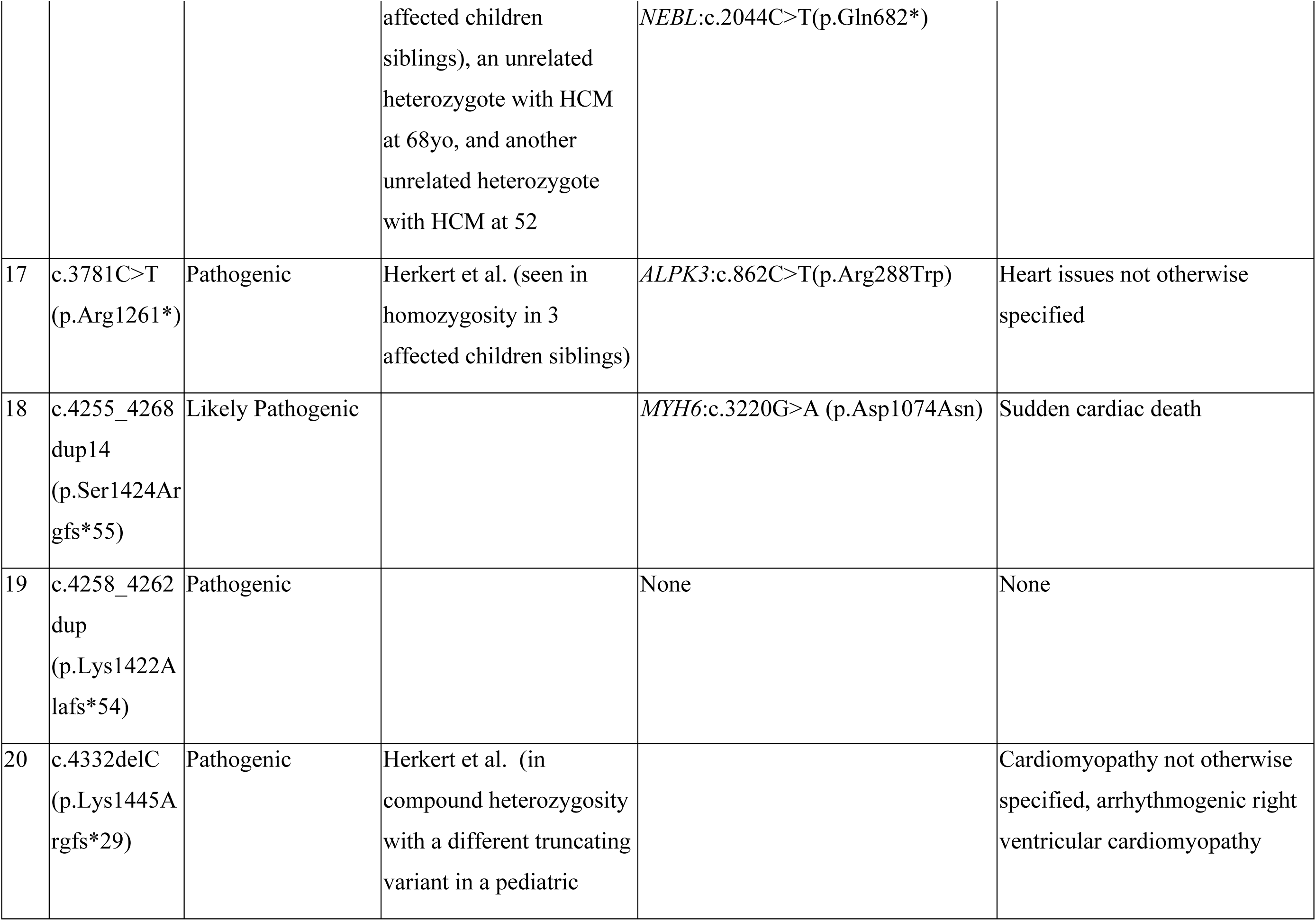
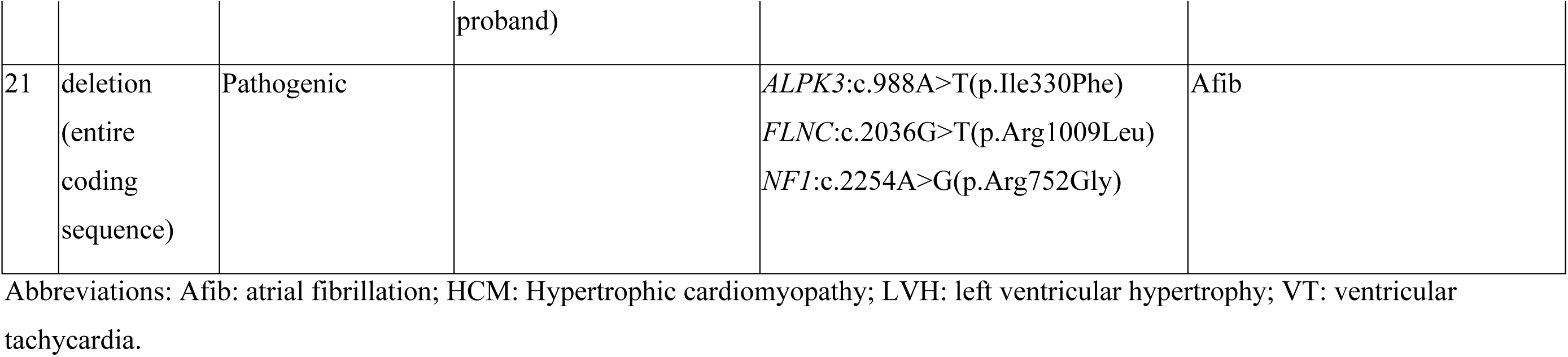
Genotypic characteristics of individuals with heterozygous *ALPK3*tv.

**Table 2.**
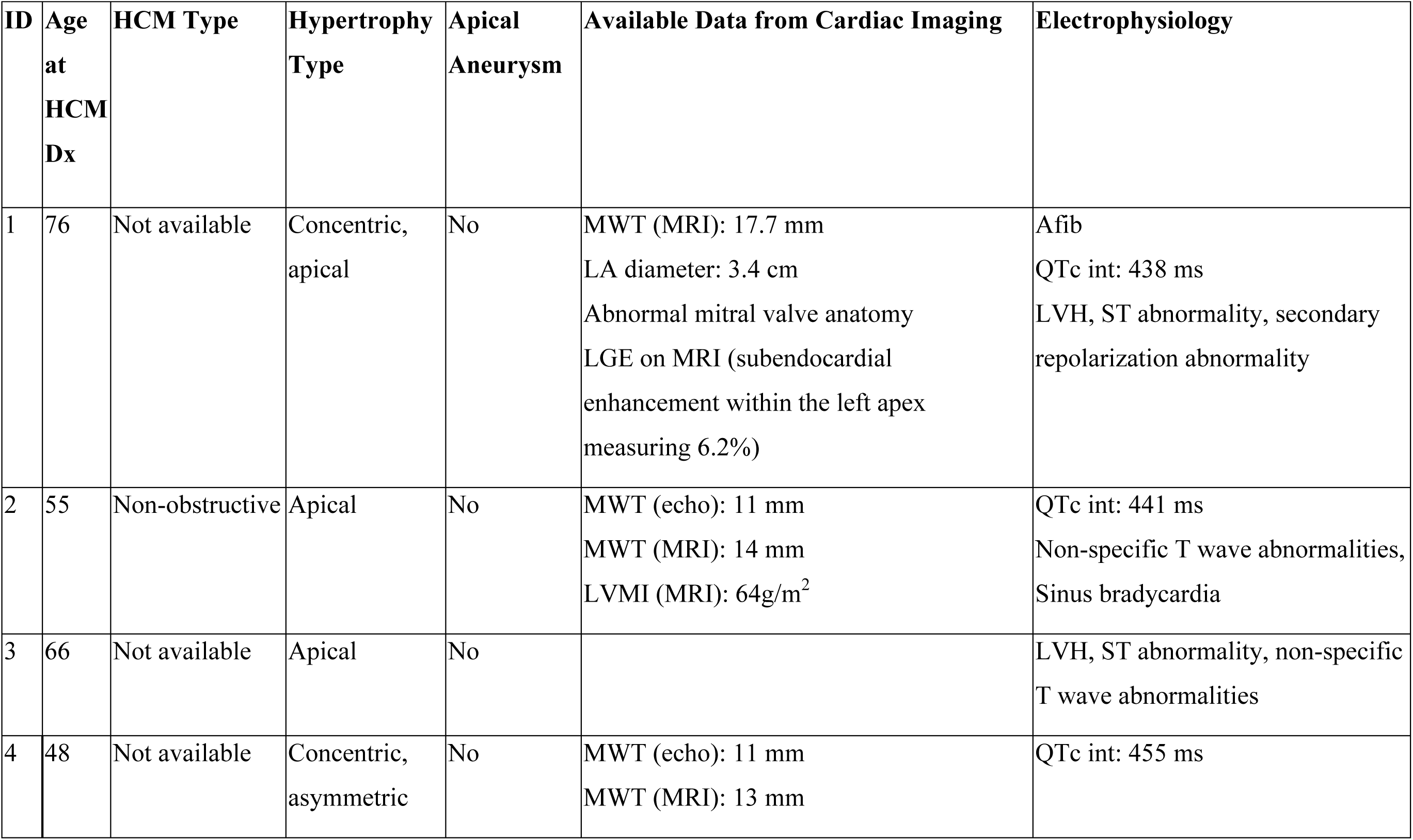
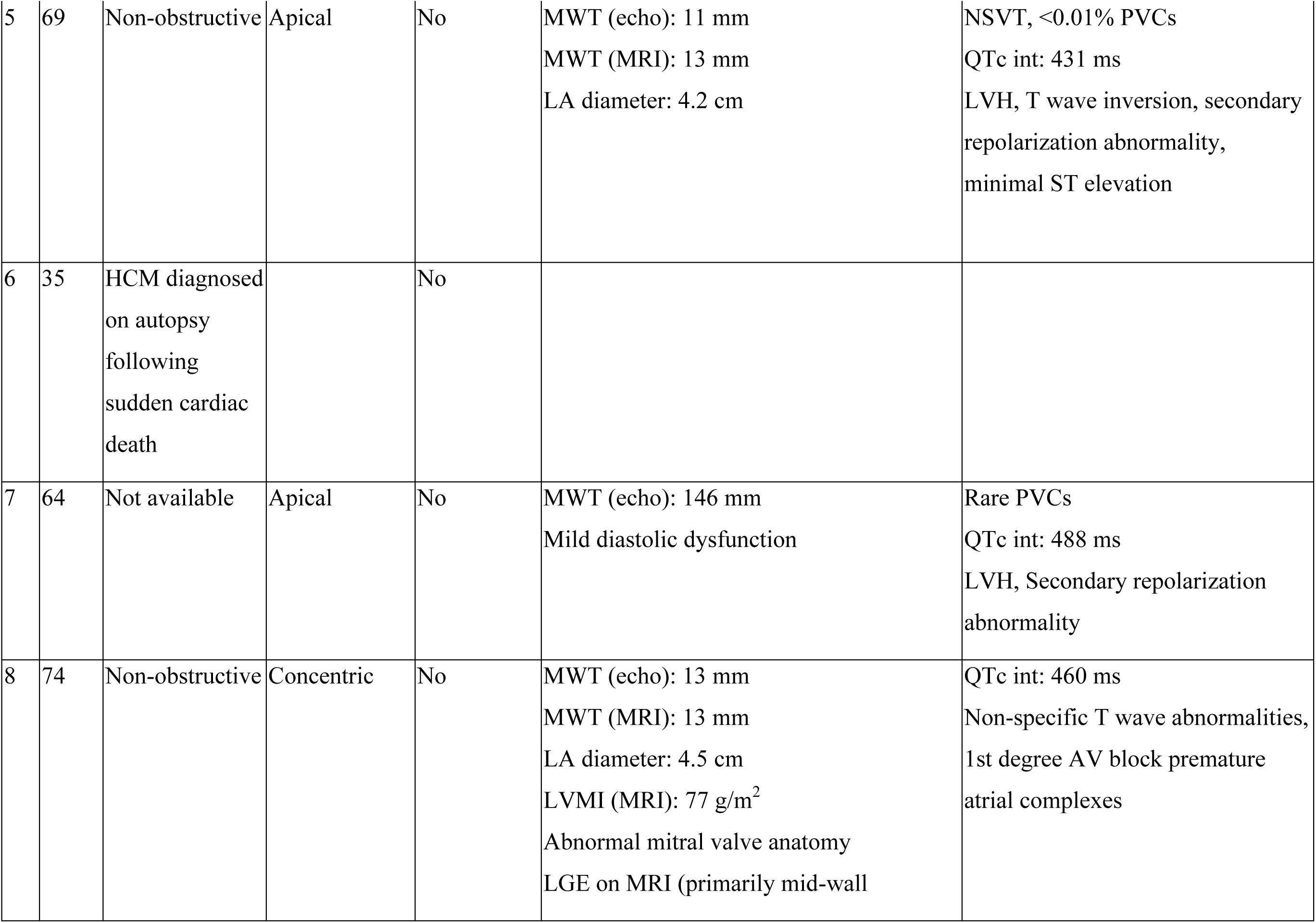
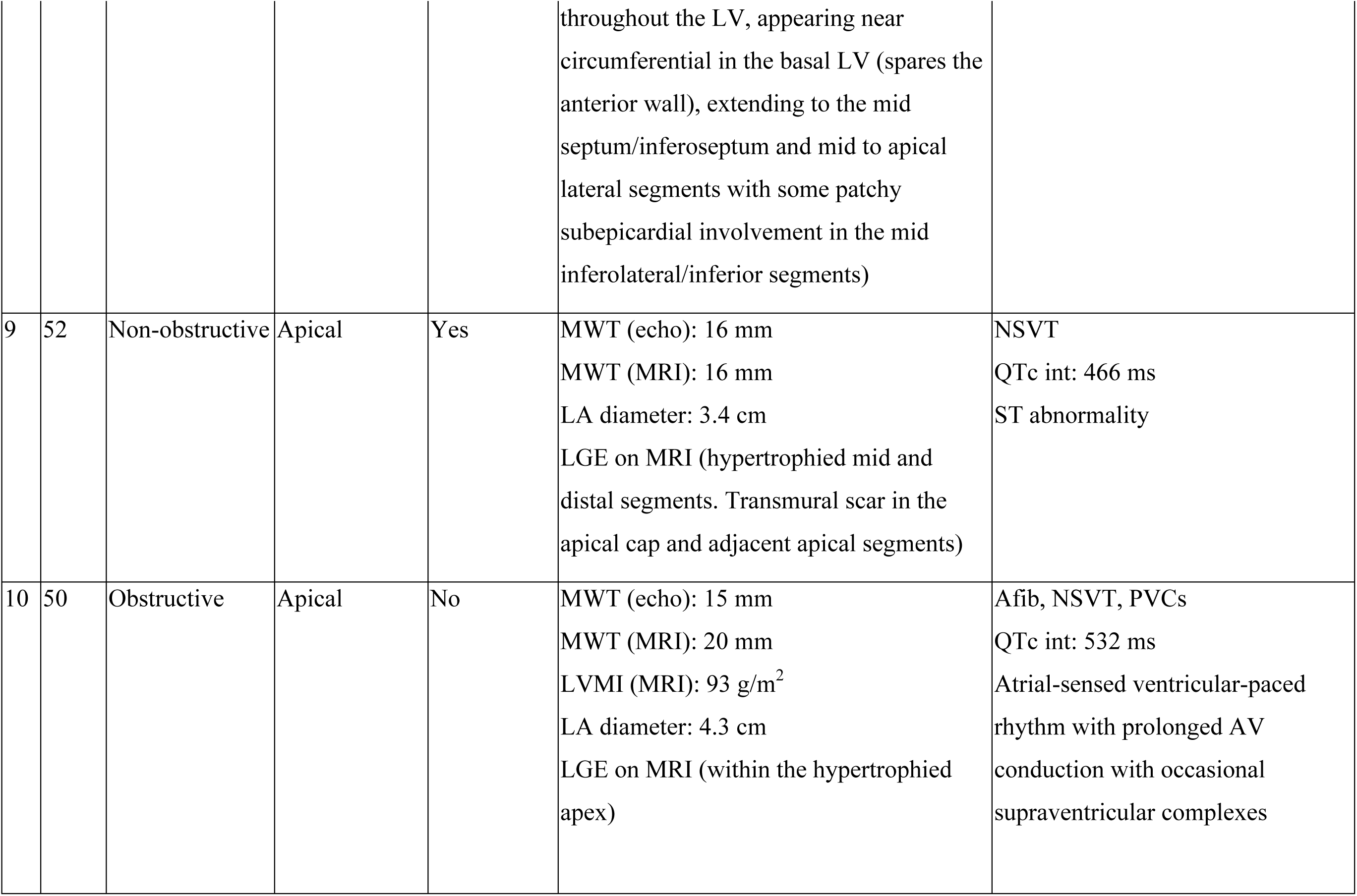
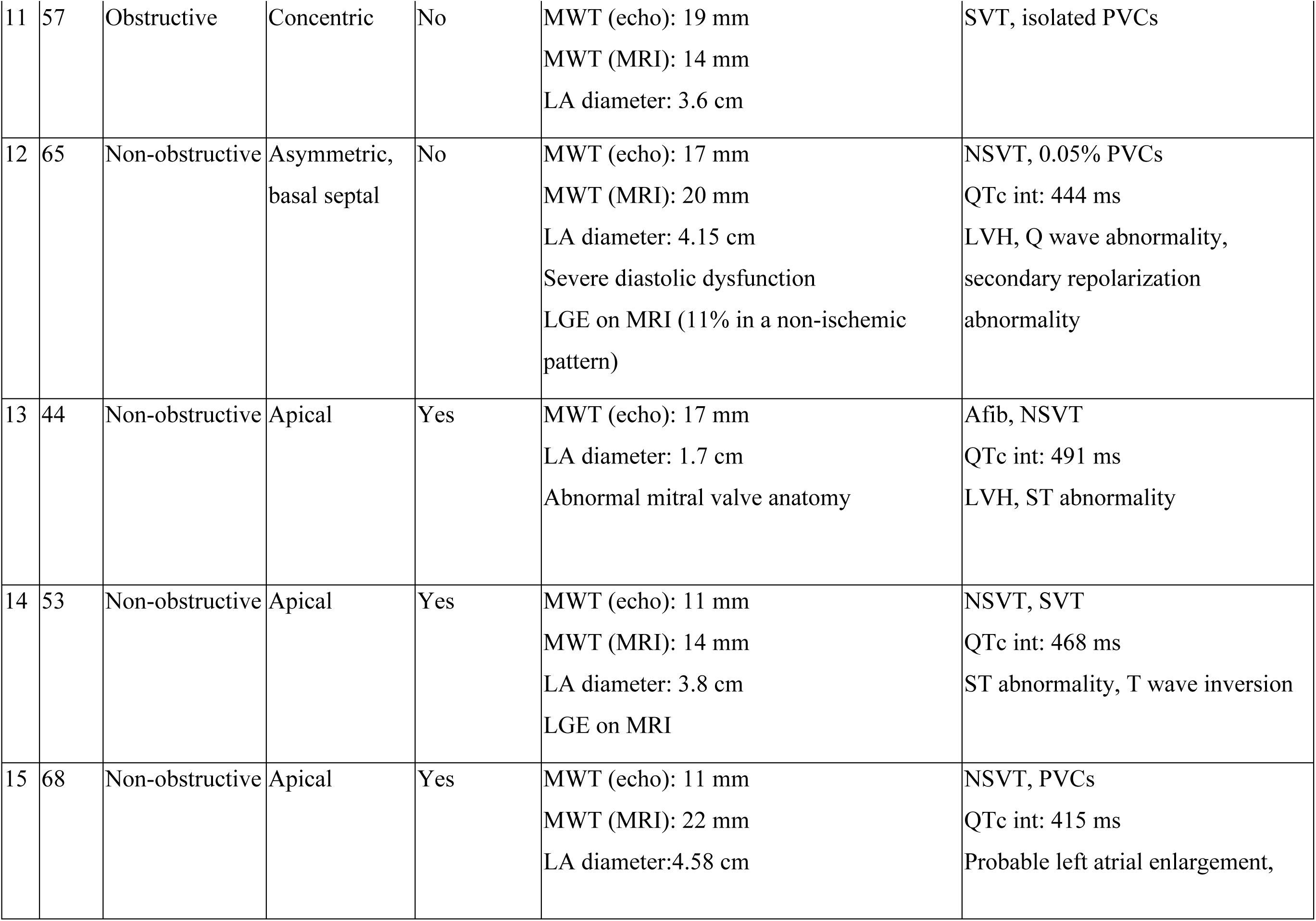
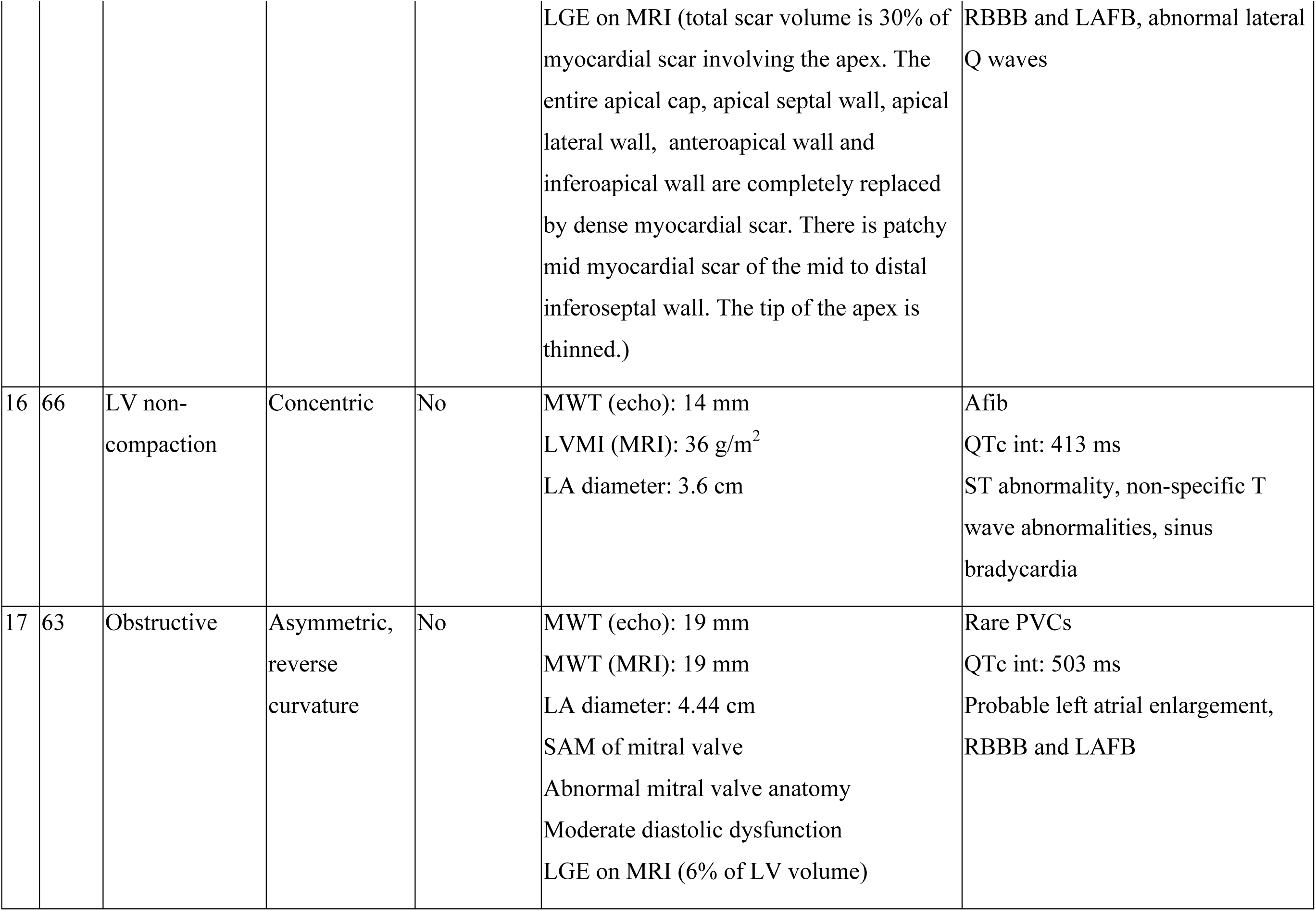
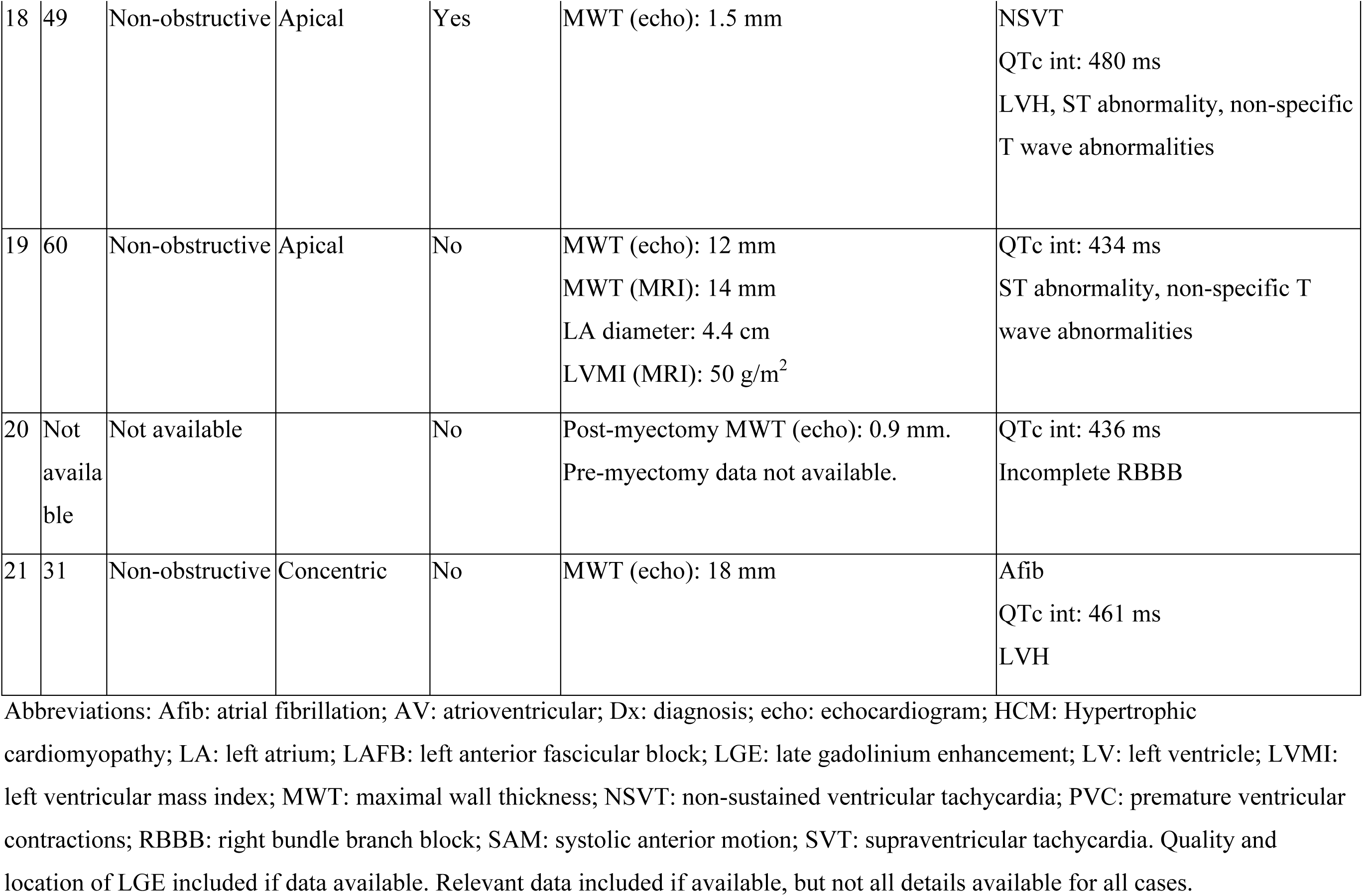
Phenotypic characteristics of individuals with heterozygous *ALPK3*tv.

Sixteen different variants spanning *ALPK3* were observed, including 12 that have not been previously reported (Figure 1). The c.2850dup (p.Ala951Serfs*6) variant recurred in 3 patients - two affected siblings (subject 10 and 11) and one presumably unrelated individual. The c.903del (p.Ile301Metfs*10), c.1018C>T (p.Gln340*), c.1490dup/c.1491C>A (p.Tyr497*), and c.3781C>T (p.Arg1261*) variants were each observed twice in our cohort in presumably unrelated families. We observed one deletion that spanned the entire *ALPK3* gene.

**Figure 1.**
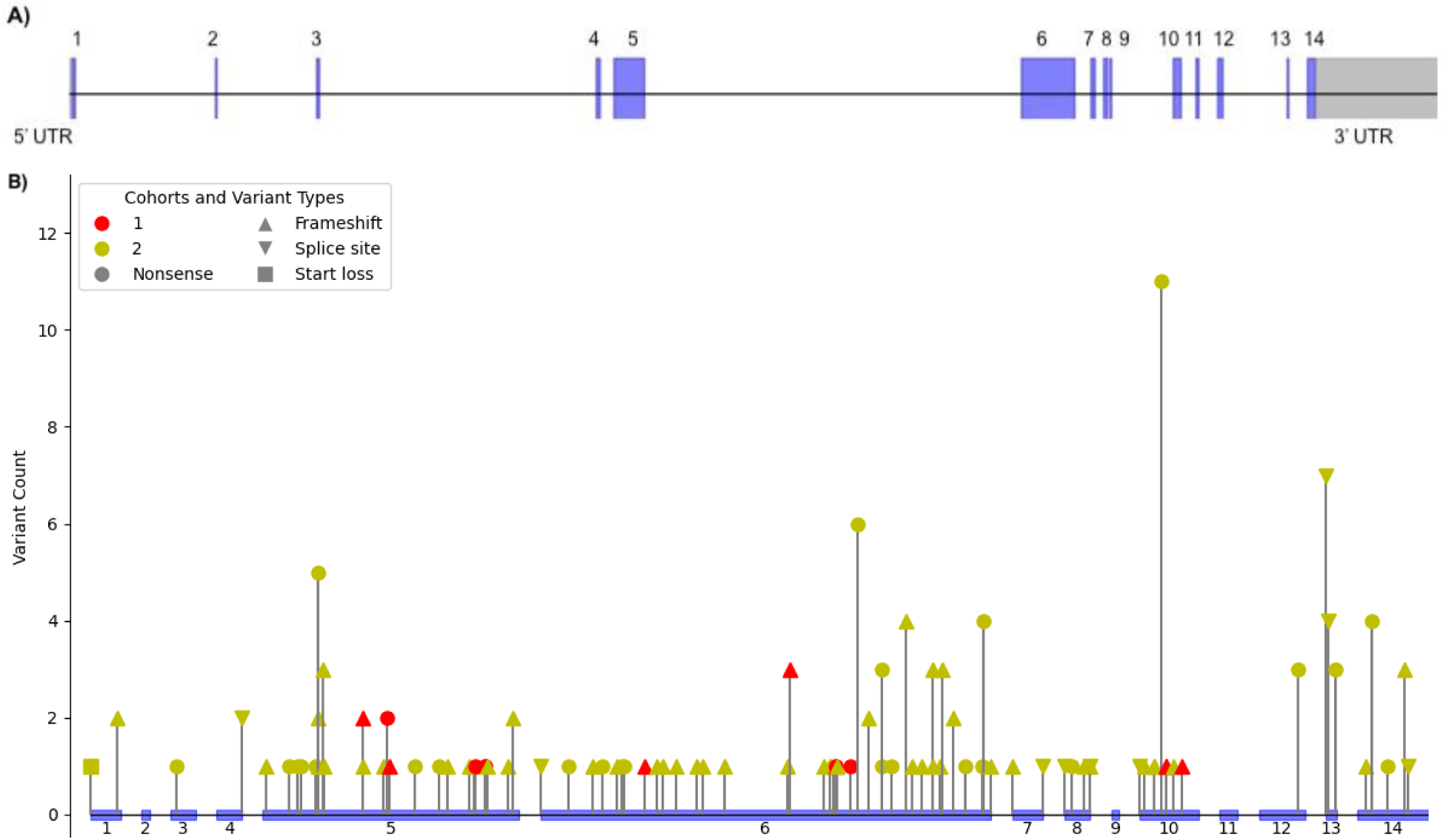
A) Schematic of ALPK3 gene. Exons 1-14 are represented in blue and untranslated regions (UTRs) are represented in grey. **B) Distribution of *ALPK3tv* associated with HCM or LVH along the gene.** Variants seen in our cohort are indicated in red. Previously reported ALPK3tv associated with HCM or LVH are indicated in yellow. The size of the exons relative to introns is not to scale.

Detailed family history was provided by the clinical genetic counselor involved in the patient’s care. Figure 2 shows the 5/20 families with significant family history with a pedigree available; 2 individuals (10%) had family history of cardiomyopathy (1 HCM, 1 cardiomyopathy of unspecified type), and an additional 4 individuals (20%) had family history of diagnoses potentially related to HCM (2 sudden cardiac death, 1 heart transplant, 1 LVH and VT (pedigree not shown)). Additional clinical details are summarized in Table 2 and available ECG tracings are shown in Figure 3.

**Figure 2:**
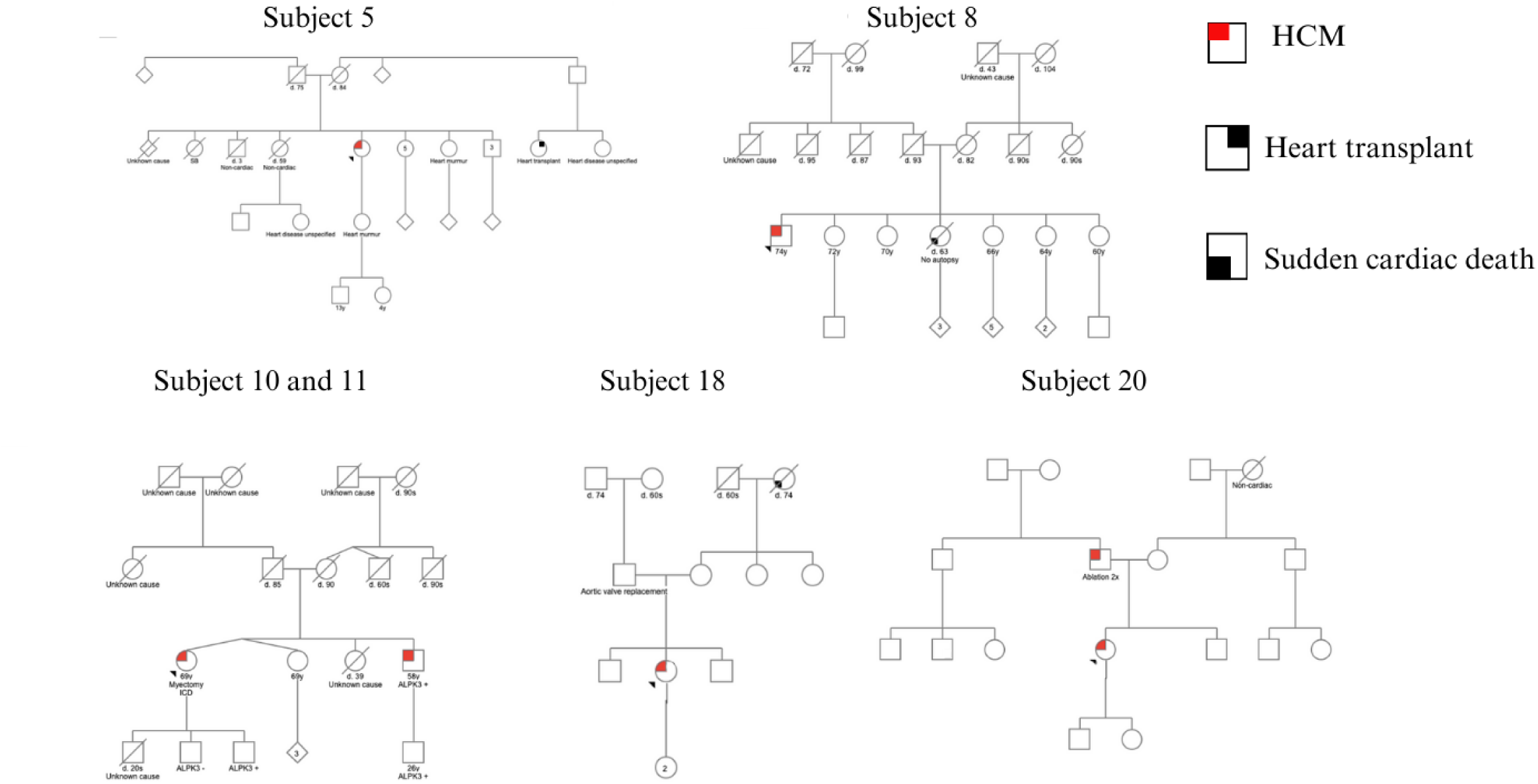
Pedigrees of ALPK3tv individuals with family history of HCM or diagnoses potentially related to HCM

**Figure 3:**
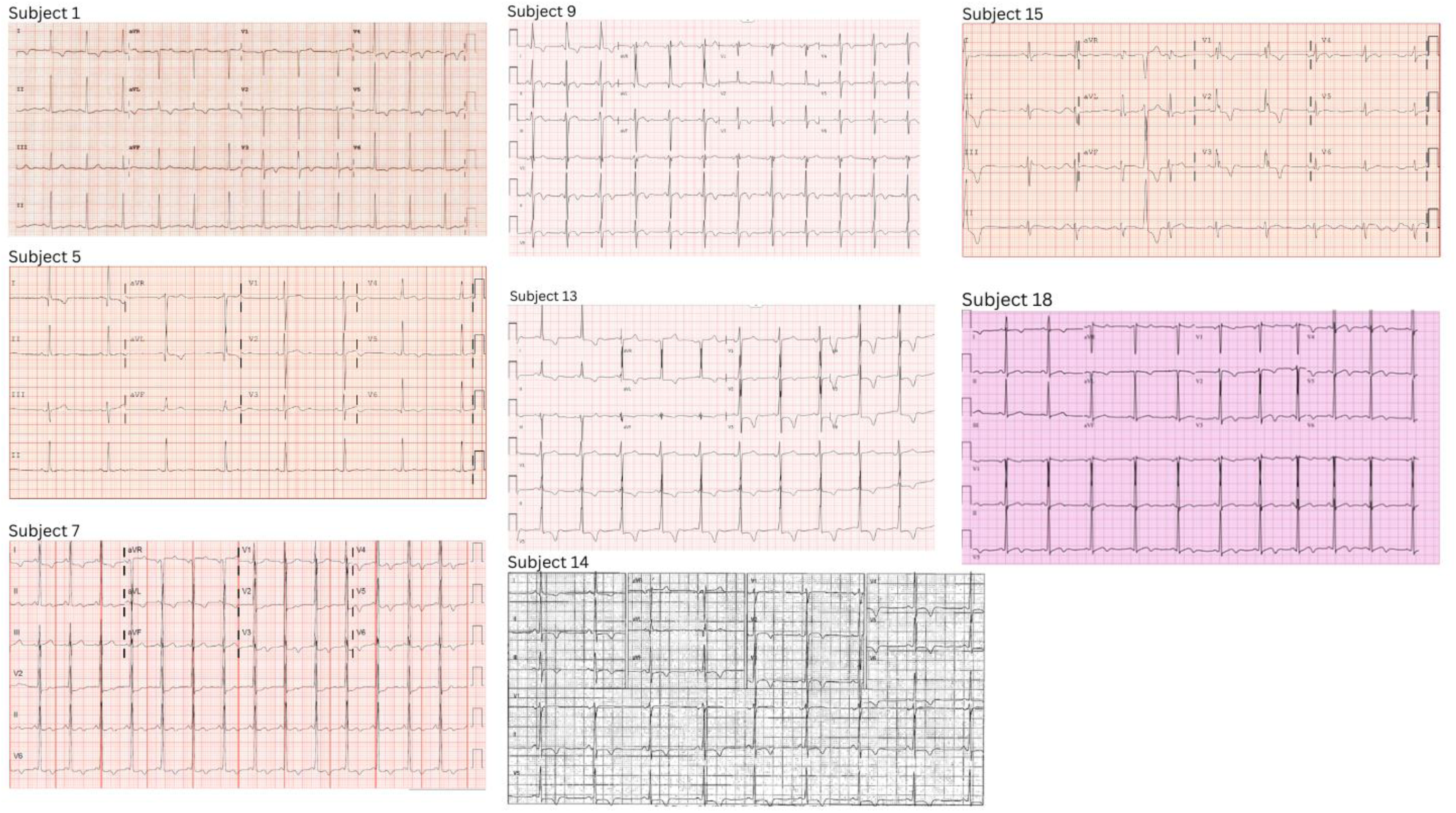
ECGs of ALPK3tv cases

### Characterization of comparison cohort (sarcomeric HCM)

From an existing clinical database of cardiovascular genetics patients at UCSF, all 132 with a clinical diagnosis of HCM and a pathogenic (116, 88%) or likely pathogenic (16, 12%) variant in a sarcomeric gene *MYBPC3, MYH7, MYL2, MYL3, ACTC1, TPM1, TNNT2, TNNC1, TNNI3,* or *ACTN2* were identified and included as a comparison group. The comparison group was similar to the ALPK3tv group with regard to the presence of hypertension and other comorbidities (see Table 3). 90% of patients in the comparison group were evaluated by an HCM specialist at UCSF. The patients underwent genetic testing between 2008 and 2024 predominantly via Invitae Laboratory (82/132, 62%) or GeneDx (35/132, 27%). Approximately half (65/132, 49%) had deleterious variants in *MYBPC3*, and the remainder had deleterious variants in *MYH7* (45/132, 34%), *TNNT2* (7/132, 5%), *MYL3* (6/132, 5%), *TNNI3* (6/132, 5%), *TPM1* (2/132, 2%), and *TNNC1* (1/132, 1%). These variants were primarily identified on HCM panels (77/132, 58%), combined cardiomyopathy and arrhythmia panels (28/132, 21%), cardiomyopathy panels (10/132, 8%), or through familial variant testing (12/132, 9%). 53% (70/132) had a family history of HCM, and an additional 24% (32/132) had a family history of cardiac arrest or atrial fibrillation.

**Table 3.**
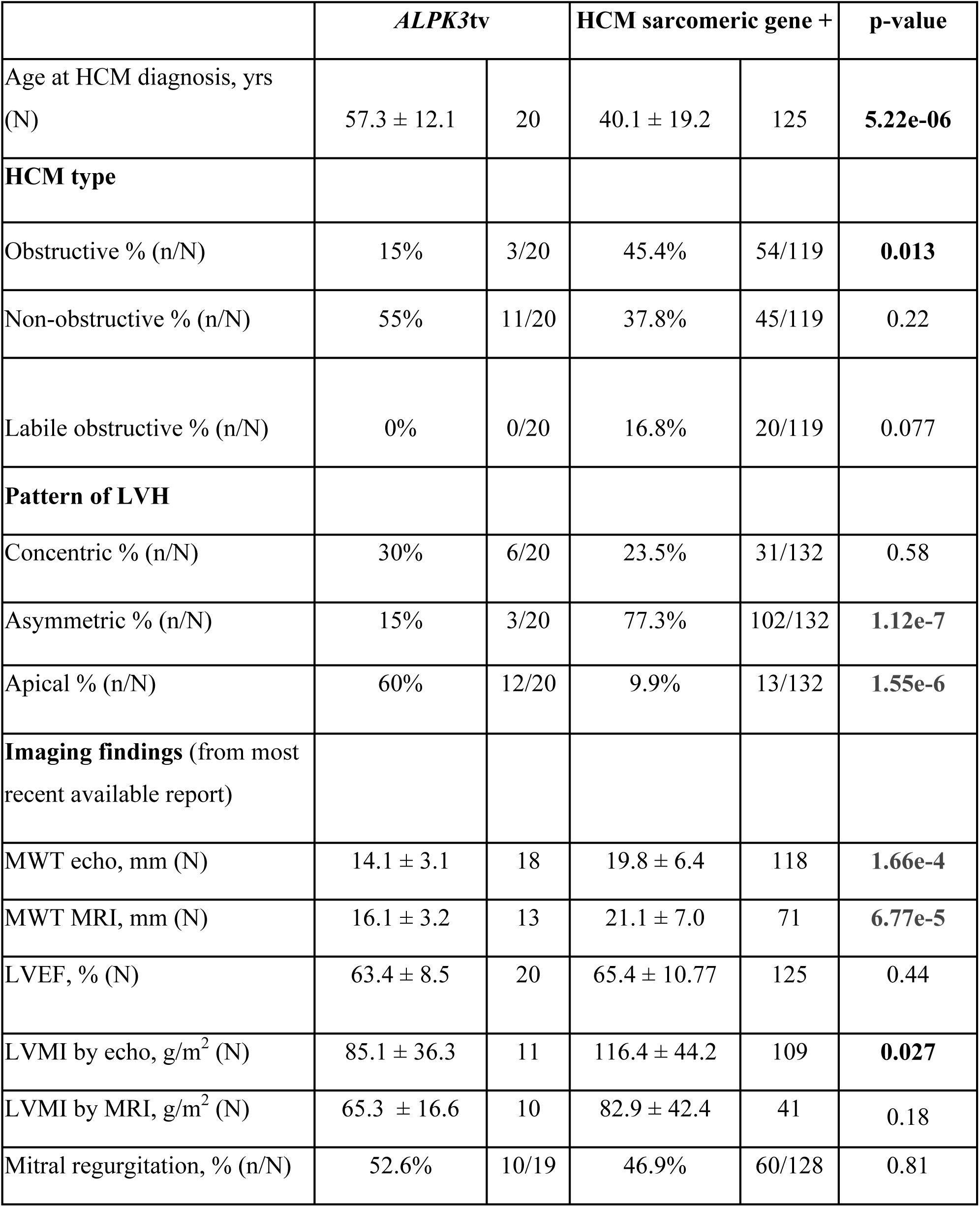
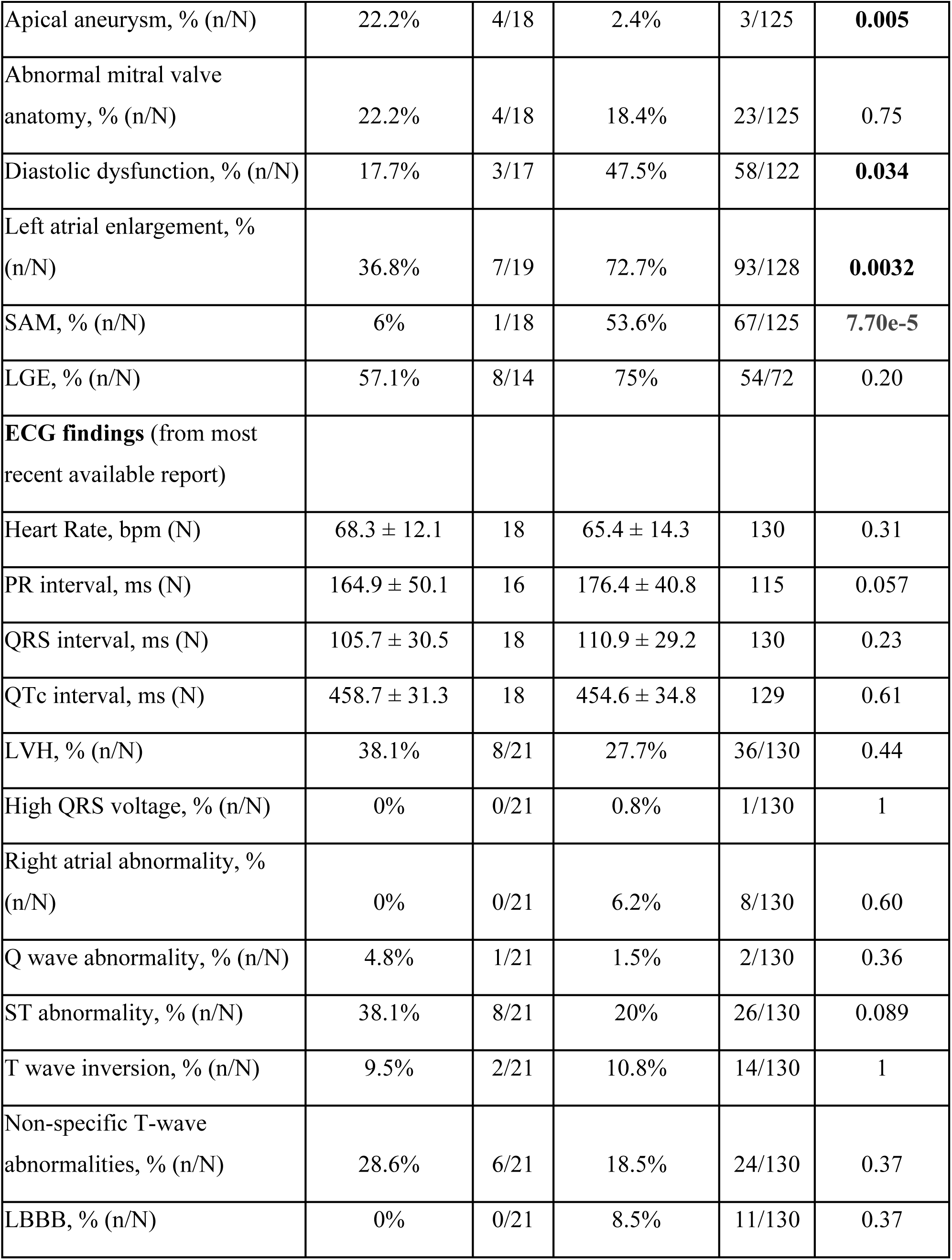
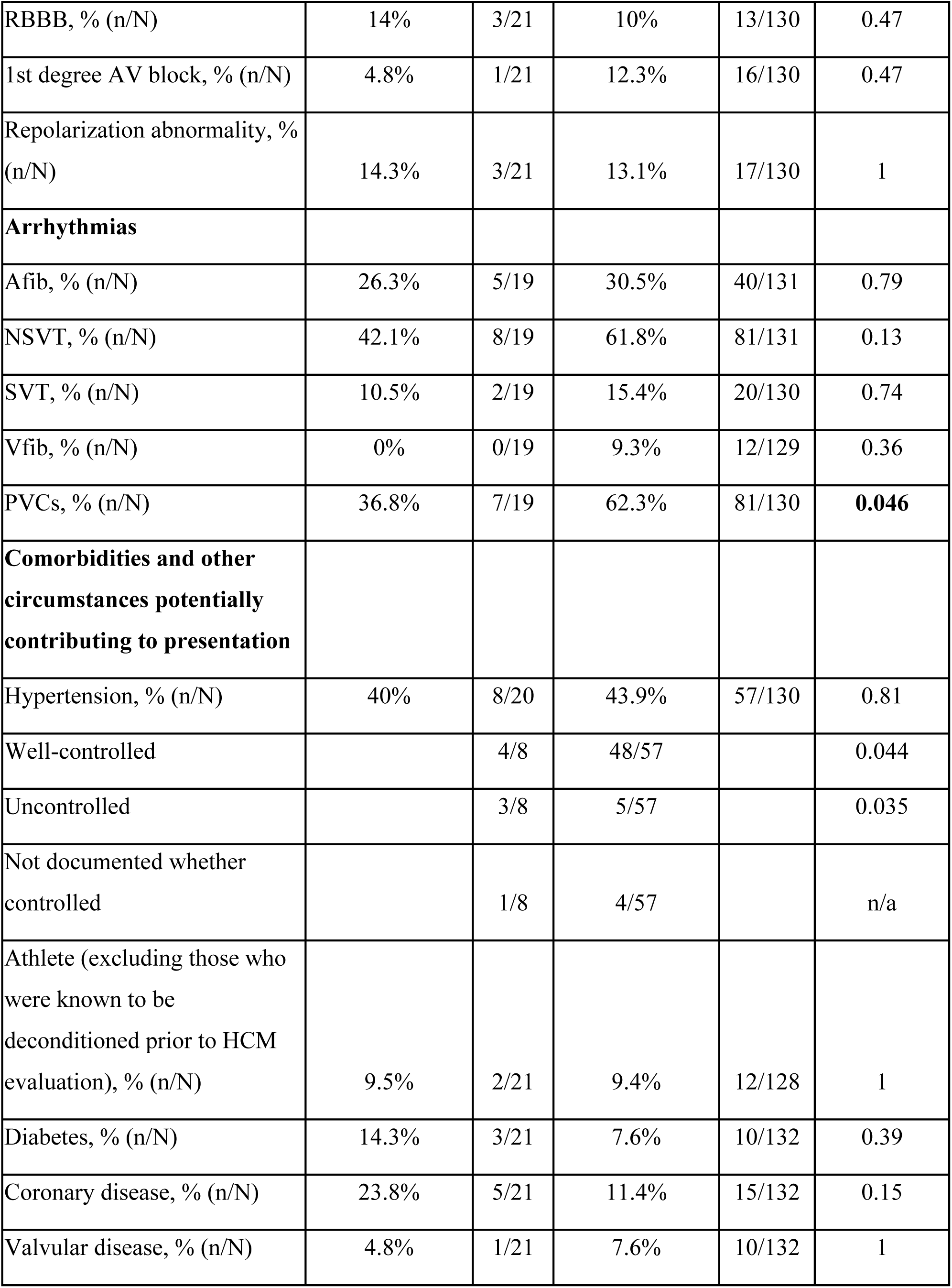
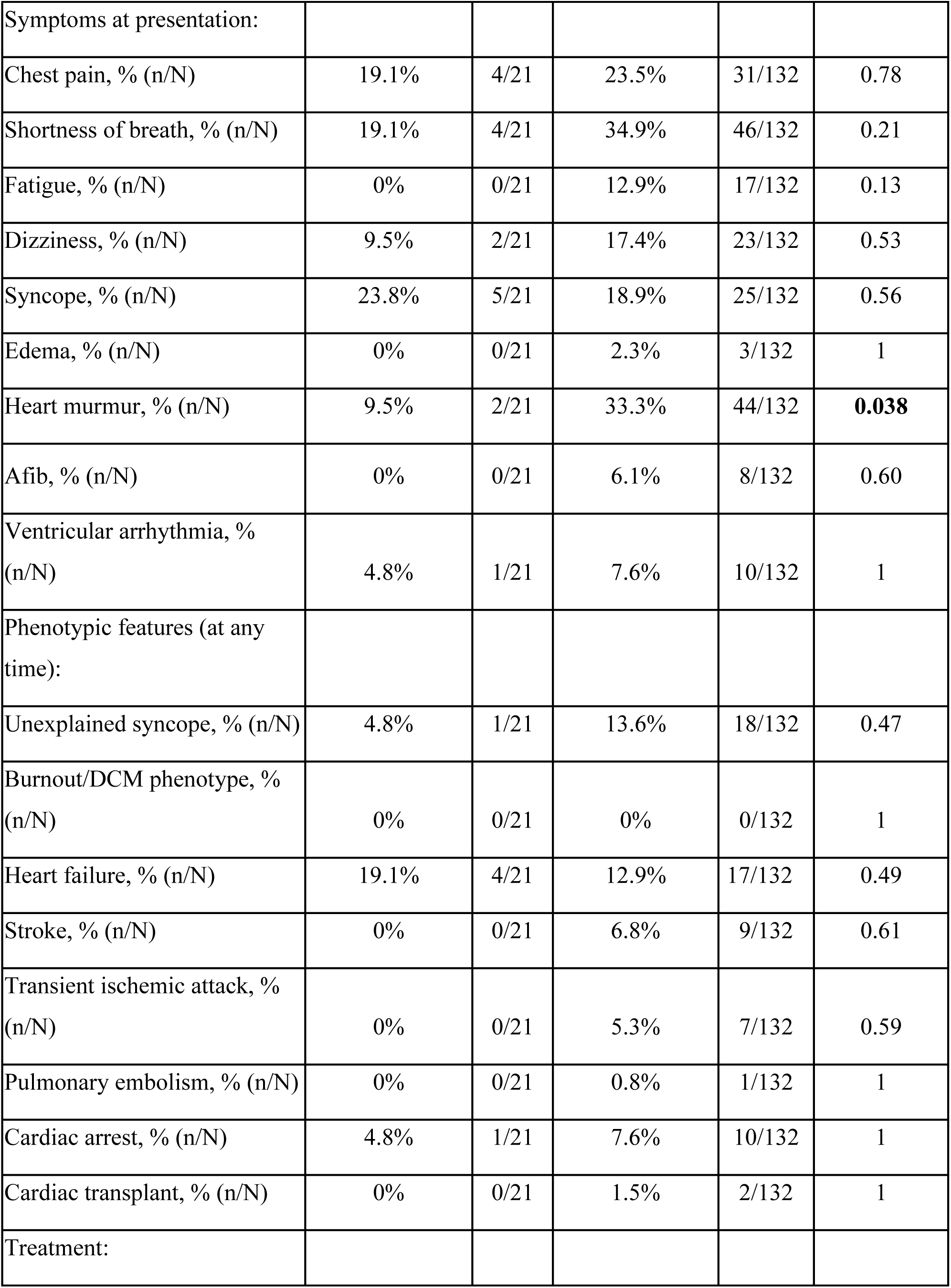
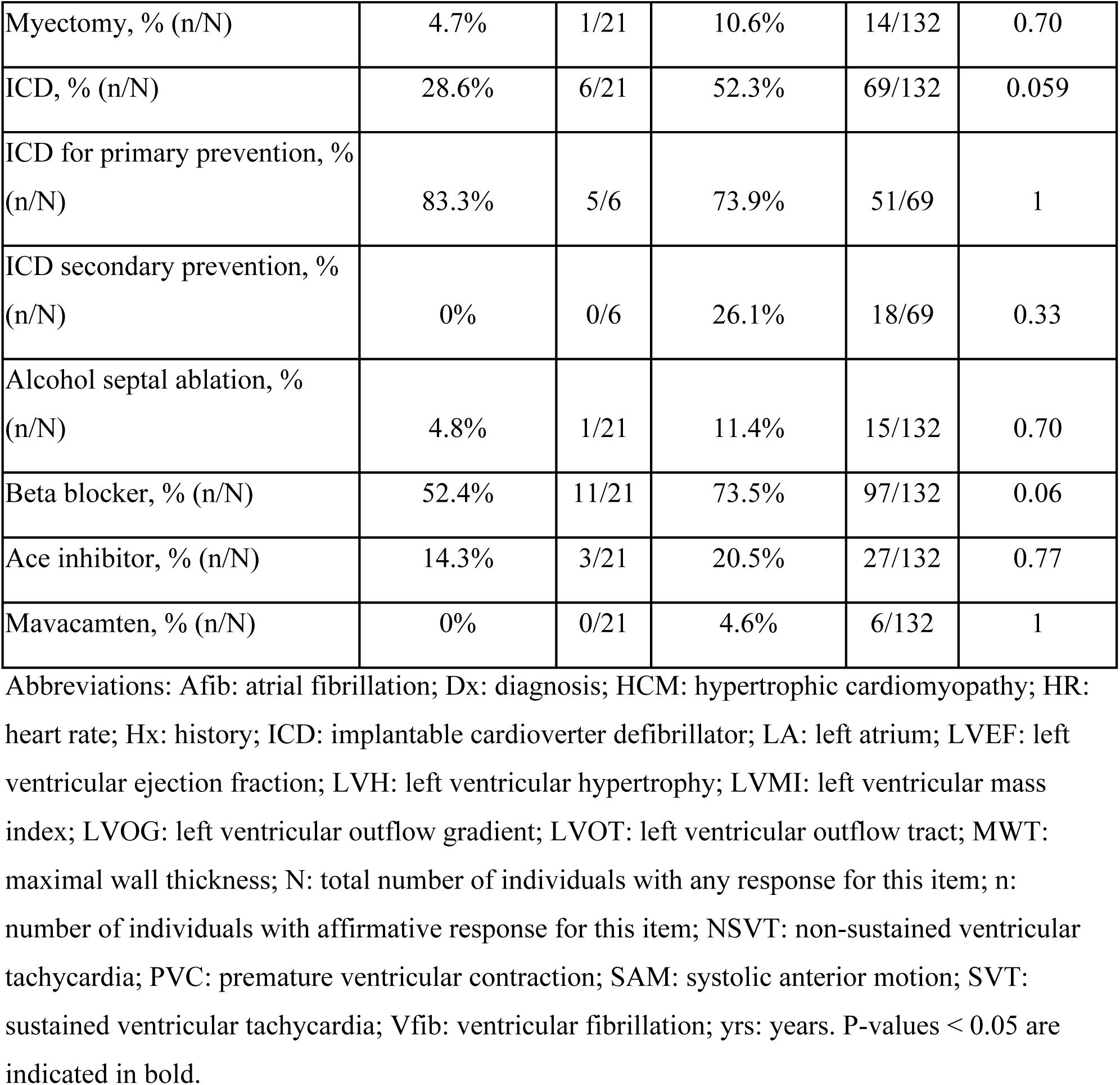
Clinical characteristics of individuals with heterozygous *ALPK3*tv and with sarcomeric genetic forms of HCM.

### Clinical Characteristics of ALPK3-related HCM compared to sarcomeric HCM

The clinical characteristics of the ALPK3tv group and the sarcomeric comparison group are summarized in Table 3. HCM caused by ALPK3tv presented later in life (57.3 ± 12.1 years vs 40.1 ±19.2 years; p = 5.22e-06) and with lower left ventricular maximal wall thickness (MWT) measured by both echocardiogram (14.4 ± 2.9 mm vs 19.8 ± 6.4 mm, p=1.66e-4) and cardiac MRI (16.1 ± 3.2 mm vs 21.1 ± 7.0 mm, p=6.77e-5). The ALPK3tv group had lower left ventricular mass index measured by echocardiogram (85.1 ± 36.3 g/m^2^ vs 116.4 ± 44.2 g/m^2^, p = 0.027) and by MRI, although the latter was not statistically significant (65.3 ± 16.6 g/m^2^ vs 82.9 ± 42.4 g/m^2^, p=0.18). The ALPK3tv group also had less left atrial enlargement than the sarcomeric group (36.8% vs 72.7%, p=0.0032). Obstructive HCM and systolic anterior motion of the mitral valve were less common in the ALPK3tv group (15% vs 45.4%, p=0.013; 6% vs 53.6%, p=7.70e-5 respectively). Diastolic dysfunction was also less common in the ALPK3tv group (17.7% vs 47.5%, p=0.034). Over half (60%, 12/20) of individuals with ALPK3tv had an apical pattern of LVH, whereas only 9.9% (13/133) of individuals with sarcomeric variants had apical LVH (p=1.55e-6).

Significantly fewer individuals in the ALPK3tv group had PVCs (36.8% vs 62.3%, p=0.046), although other ventricular and atrial arrhythmias were similar in each group. Although one individual with ALPK3tv had a fatal cardiac arrest at 35 years old, no significant difference was seen in the rate of ventricular fibrillation. Apart from a shorter PR interval in the ALPK3tv of borderline significance (164.9 ± 50.1 ms vs 176.4 ± 40.8 ms, p=0.057), other ECG intervals were similar between the two groups.

Apical aneurysm was relatively common in the ALPK3tv group although rare in the sarcomeric group (22.2% vs 2.4%, p=0.005). The older age of diagnosis of HCM in the ALPK3tv did not seem to explain the higher proportion of apical aneurysm; the age of diagnosis of HCM in ALPK3tv and sarcomeric patients with apical aneurysm was not significantly different (53.3 ± 10.4 years for ALPK3 vs 37.3 ± 20.4 years for sarcomeric, p=0.4). All four of the ALPK3tv cases with apical aneurysms had non-obstructive HCM with apical hypertrophy. 2/3 of the sarcomeric HCM cases with apical aneurysms had obstructive HCM one had labile- obstructive HCM, and all had asymmetric LVH pattern.

Significantly fewer individuals in the ALPK3tv group had PVCs (36.8% vs 62.3%, p=0.046), although other arrhythmias were similar in each group. No ventricular fibrillation was seen in the ALPK3tv group although it was not common in the sarcomeric group either. ECG findings were similar in the two groups; the PR interval was non-statistically significantly shorter in the ALPK3tv group (164.9 ± 50.1 ms vs 176.4 ± 40.8 ms, p=0.057).

## DISCUSSION

Here we describe a new cohort of 20 unrelated individuals with heterozygous ALPK3tv HCM and offer a detailed phenotypic comparison with HCM patients harboring deleterious variants in sarcomeric genes. Similar to previous reports, we find later onset disease, although we do find, for the first time, a higher-than-expected prevalence of apical aneurysm in ALPK3tv patients. This finding may have important genotype-specific clinical implications.

### Comparison to Other ALPK3tv Cohorts

Lopes et al. also found a later age of onset in ALPK3tv (56 ± 15.9 years) compared to individuals with sarcomeric HCM ^14^, while Ader et al. reported a similar age of onset for ALPK3tv (53 ± 16.9 years) ^15^ as our cohort (57.3 ± 12.1 years). In contrast, Wang et al. ^16^ and Ryu et al. ^17^ reported younger age of onset (40.8 ± 16.1 years and 42 ± 11.4 years, respectively), although Ryu et al. ^17^ used broader inclusion criteria, and Wang et al. included individuals in their ALPK3tv cohort who also had a pathogenic variant in a sarcomeric gene (n=2, 11%) ^16^. In contrast to Lopes et al. ^14^ and Wang et al. ^16^ who showed no significant difference in MWT in ALPK3tv HCM compared to sarcomeric HCM, our ALPK3tv cohort had significantly smaller MWT measurements on echocardiogram than both our own comparison group and the ALPK3tv patients from the prior studies. We are the first to report MWT measurements from cardiac MRI. Cardiac MRI may more accurately measure MWT than echocardiogram ^19^. When compared to our sarcomeric HCM cohort, our ALPK3tv cohort also had significantly smaller MWT measurements on cardiac MRI. LV mass index from MRI rather than echocardiogram may be useful in quantifying the degree of hypertrophy in HCM patients; our data show a trend towards lower LV mass index in the ALPK3tv group although the difference did not reach statistical significance.

### Patterns of LVH in ALPK3tv HCM

Apical LVH was seen in more than half of the patients with ALPK3tv, and only 10% of the sarcomeric group. Apical LVH was similarly prevalent in ALPK3 HCM in other studies ^14,16,17^. Published figures estimate apical HCM accounts for a quarter of HCM cases in Asian populations and 1-10% in non-Asian populations ^20^. The prevalence of apical morphology in ALPK3 is clinically relevant not only because apical HCM is typically associated with lower mortality risk ^21,22^ but it may also influence selection of imaging modality. Cardiac MRI is better suited than echocardiogram for visualizing apical HCM ^23,24^. Obstructive HCM was uncommon in the ALPK3tv group. In line with that, systolic anterior motion of the mitral valve (SAM) was only seen in 1/18 individuals (6%) in the ALPK3tv group, whereas more than half of the sarcomeric group had SAM (53.6%, p=7.70e-5). We also observed a lower rate of diastolic dysfunction in the ALPK3tv group when compared to sarcomeric HCM. Diastolic dysfunction is associated with a worse HCM prognosis independent of left ventricular outflow tract obstruction^19^.

### Prevalence of Apical Aneurysm in ALPK3tv HCM

This is the first study to report a significant enrichment for LV apical aneurysms in individuals with ALPK3tv. While apical aneurysm is estimated to occur in 2% of those with HCM and 13-15% of those with apical HCM ^21^, 22.2% of our ALPK3tv HCM cohort and 33.3% of those with ALPK3tv with apical hypertrophy had apical aneurysms. In contrast, only 2.4% of the sarcomeric HCM group and 0% of those with sarcomeric apical HCM had apical aneurysms. Apical aneurysms in HCM are thought to arise from a variety of mechanisms including mid ventricular obstruction and auto-infarction in hypertrophied regions ^25^. Considering that none of the patients with ALPK3tv and apical aneurysms in our cohort had obstructive HCM, this suggests that apical aneurysms in the absence of obstructive HCM or other cause may increase clinical suspicion of an ALPK3tv. LGE data was available for 2 of the 4 ALPK3tv cases with apical aneurysm; LGE was present in both, one case with large scar volume of 30% involving the apex, and the other had LGE present but further details were not available. Further studies of LGE quantification and distribution in patients with LV apical aneurysm in the setting of ALPK3tv would be valuable to elucidate the mechanism of apical aneurysm development in these patients.

### Penetrance of ALPK3tv HCM

The ALPK3tv group had less family history than the control HCM group, suggesting lower penetrance, similar to findings previously reported ^16^. Here we include 3 generation pedigrees (Figure 2). However, to assess penetrance more robustly, further studies should include clinical evaluation of relatives or potentially interrogate large genetic databases with population-level clinical and genetic data.

### Implications for Clinical Management

The current American Heart Association and American College of Cardiology clinical practice guideline includes class 1 evidence for cardiac MRI being beneficial in patients with HCM who are not otherwise identified as high risk for sudden cardiac death to assess for features including apical aneurysm ^26^. Given the prevalence of apical aneurysm and otherwise lack of high risk features for sudden cardiac death in ALPK3tv HCM, cardiac MRI may be important in individuals with ALPK3tv genotype in order to accurately stratify their risk. Longitudinal assessment of these patients using contrast echo and MRI may also be beneficial for assessing disease progression with respect to apical hypertrophy, LGE and apical aneurysm. Moreover, apical aneurysm in HCM confers an increased risk of thromboembolism, which may warrant consideration of prophylactic anticoagulation in these patients ^27^.

## LIMITATIONS

Some data were unavailable for review in the medical record, as documentation practices may vary between institutions and some cardiac investigations may have been completed outside of the primary institution. Chart review was performed at a single point in time, and additional clinical features may have become apparent after that time.

Patients with apical HCM in some cases may have hypertrophy purely in the apex and therefore standard echocardiography may not detect this. We infer that most of the ALPK3tv cases included in this study did have appropriate contrast echocardiogram and measurement of the apex as 75% of cases were collected from institutions affiliated with HCM Centers of Excellent. However, some echocardiogram measurements of hypertrophy may be underestimated. We did not collect data on LV cavity dimension. Future studies further characterizing the apical hypertrophy and implications for LV cavity size and stroke volume would be informative.

## CONCLUSION

Heterozygous ALPK3tv HCM causes incompletely penetrant late onset HCM that is associated with lower wall thickness, lower rate of diastolic dysfunction, and less frequent left atrial enlargement than HCM caused by deleterious variants in sarcomeric genes. Findings from our cohort suggest that individuals with ALPK3tv HCM have higher rates of apical hypertrophy and apical aneurysm.

## Data Availability

The data that support the findings of this study are available from the corresponding author upon reasonable request.

## ACKNOWLEDGEMENTS

We would like to thank James Pirruccello, MD, PhD, for his helpful discussions and critical review of the manuscript, and Sophia Sussman, genetic counseling graduate student, for her help proof-reading the manuscript.

## SUPPLEMENTARY MATERIAL

Table S1

## Non-standard Abbreviations and Acronyms

Afib: atrial fibrillation
AV: atrioventricular
CGC: certified genetic counselor
Dx: diagnosis
ECG: electrocardiogram
Echo: echocardiogram
HCM: hypertrophic cardiomyopathy
HR: heart rat
Hx: history
ICD: implantable cardioverter defibrillator
LA: left atrium
LAFB: left anterior fascicular block
LGE: late gadolinium enhancement
LV: left ventricle
LVEF: left ventricular ejection fraction
LVH: left ventricular hypertrophy
LVMI: left ventricular mass index
LVOG: left ventricular outflow gradient
LVOT: left ventricular outflow tract
MWT: maximal wall thickness
NSVT: non-sustained ventricular tachycardia
PVC: premature ventricular contractions
RBBB: right bundle branch block
SAM: systolic anterior motion
SIG: special interest group
SVT: supraventricular tachycardia
tv: truncating variant
UCSF: University of California, San Francisco
Vfib: ventricular fibrillation
VT: ventricular tachycardia

